# Epigenetic age acceleration in offspring linked to paternal smoking initiation and overweight in puberty: Evidence from a two-generation study

**DOI:** 10.64898/2026.05.05.26352444

**Authors:** Juan P López-Cervantes, Toril Mørkve Østergaard, Negusse T. Kitaba, Marianne Lønnebotn, Randi J. Bertelsen, Simone Accordini, Christer Janson, Shyamali C Dharmage, Karl A Franklin, Francisco Javier Callejas González, Mathias Holm, Ane Johannessen, Caroline Lodge, Andrei Malinovschi, Anna Oudin, Francisco Gómez Real, Anders Flataker Viken, Vivi Schlünssen, John W. Holloway, Cecilie Svanes

## Abstract

**Background:** Father’s adolescent smoking and overweight affect respiratory health in offspring, suggesting that paternal puberty exposures may influence offspring biological ageing through preconception epigenetic mechanisms.

**Methods:** We analyzed epigenetic age acceleration using four validated epigenetic clocks derived from blood DNA methylation in 892 RHINESSA offspring (mean age 27 years), linked to parental data on smoking and body shapes from RHINE/ECRHS. Linear regression examined parental smoking initiation (≤15 or >15 years) and overweight body shape (childhood/puberty or age 30) in relation to offspring epigenetic age acceleration, adjusting for offspring sex, age and parental socioeconomic status. Sensitivity analyses accounted for offspring smoking and BMI.

**Results:** PCHorvath (β 1.53; 95% CI 0.02, 2.9), PCGrimAge (1.21; 0.03, 2.1), DunedinPACE (0.04; −0.001, 0.1) and PCPhenoAge (1.92; −0.3, 4.2) were accelerated in daughters of fathers who started smoking ≤15 years. Likewise, PCHorvath (2.25; 1.2, 3.3), PCGrimAge (1.36; −0.2, 2.9), DunedinPACE (0.07; 0.01, 0.1) and PCPhenoAge (3.11; 1.8, 4.4) were accelerated in daughters and sons of fathers who had been overweight in childhood and puberty. These results remained largely unchanged after additional adjustments or stratification in sensitivity analyses. No associations were found for maternal smoking or overweight in puberty.

**Conclusions:** Epigenetic ageing is accelerated in offspring of fathers who smoked or were overweight in puberty, independent of offspring lifestyle. These findings suggest that adolescent boys’ environment and lifestyle may be critical for next-generation health.

Figure 1.
Graphical abstract
**Legend to graphical abstract Figure**
Father’s smoking or overweight during puberty was associated with accelerated epigenetic aging in offspring (n=892), independent of the offspring’s own lifestyle. No such pattern was observed for maternal puberty exposures, or when paternal exposures occurred after puberty. Male puberty may be a critical window for next-generation health.

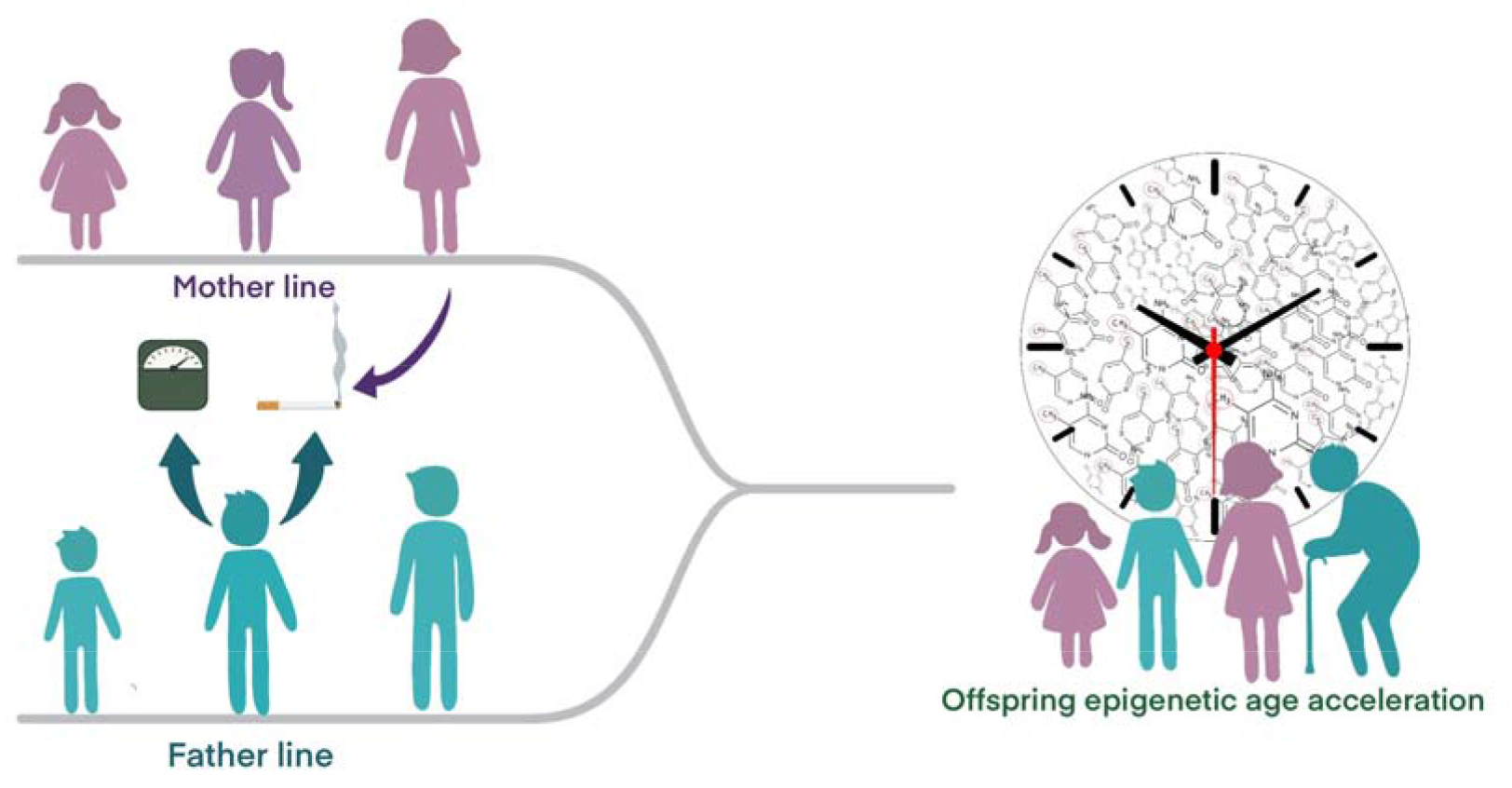

## 1. Introduction

Emerging evidence highlights the significance of parental preconception exposures in shaping offspring health. Previous studies have linked paternal smoking before age 15 and early-onset overweight to increased risk of asthma, reduced lung function and risk of obesity in children (1–6). Other exposures during parental childhood and adolescence - such as air pollution, infections, and excess food supply - further support the notion that preconception environments can influence the next generation’s respiratory, metabolic and cardiovascular health (7–9).

These findings suggest sex-specific parental effects, with paternal puberty emerging as a critical window of susceptibility, important for the health of the progeny. A preconception influence on epigenetic (re)programming during germ cell development, and a transmission and retention of altered sperm-born epigenetic marks in the developing embryo is a proposed mechanism underlying these associations (10). Notably, paternal smoking and overweight during puberty have been associated with unique DNA methylation patterns in offspring, implicating long-term impacts on metabolic and respiratory outcomes (11, 12).

Epigenetic age, derived from DNA methylation changes at specific CpGs, serves as a biomarker of biological ageing and cumulative exposures, reflecting whether a person is ageing faster than their chronological age and potentially predicting age-related diseases and mortality (13, 14). Accelerated epigenetic ageing has been linked to lifestyle factors such as smoking and high BMI (15–18) within one generation, including exposure *in utero* or early life (19). A recent paper found paternal adolescent smoking was associated with epigenetic age acceleration in cord blood (20). Further, it seems biologically plausible that exposure effects transmitted through the germline cells may influence health and disease broadly. Yet there is very limited research on the role of parental lifestyle and behaviors before conception for biological aging in future offspring.

We hypothesized that both a mixed chemical/toxic exposure - smoking, and metabolic status measured as overweight, would lead to accelerated epigenetic ageing in adult offspring, if the exposures occurred at the time of germ-cell maturation in puberty, following sex-specific patterns. Using two-generation data from the Respiratory Health in Northern Europe, Spain and Australia (RHINESSA) study, linked to parental data from Respiratory Health in Northern Europe (RHINE) and European Community Respiratory Health Survey (ECRHS) (21), we investigated whether parental smoking initiation before or after age 15 and body shapes in childhood, puberty and adulthood were associated with offspring epigenetic age acceleration, and whether these associations differed by parental and offspring sex.

## 2. Methods

### 2.1 Study design, population and data

In this two-generation analysis, we included 892 offspring (7-50 years of age) with data on blood DNA methylation from the RHINESSA baseline study (2013-16), and complete data on parental smoking/age at smoking initiation in the RHINE/ECRHS studies (offspring-father n=415; offspring-mother n=477). There was complete information on parental body shapes for 719 of these offspring (offspring-father n=338; offspring-mother n=381). The participants were from six RHINESSA study centers in Bergen, Norway; Huelva and Albacete, Spain; Melbourne, Australia; Tartu, Estonia and Aarhus, Denmark. Offspring information from the RHINESSA study was linked to their parents’ data, provided in the population-based ECRHS (www.ecrhs.org) and/or the RHINE (https://rhine.w.uib.no/) studies (21).

Ethical approval was obtained from the medical research ethics committees at each participating study center for each study and each study wave. All procedures complied with relevant ethical regulations for research involving human participants. Each participant (or legal representative) provided written informed consent.

### 2.2 Smoking and overweight in parents

Timing of smoking initiation: Defined using the responses to the questions in the ECRHS/RHINE questionnaires (1) “Are you a smoker?” (2) “Are you an ex-smoker?” If yes to (1) or (2): (3) “How old were you when you started smoking”. Thus, it was categorized as: 1) never tobacco smoking (“no” to 1 and 2; reference category), 2) tobacco smoking in puberty or ≤15 years (“yes” to 1 and/or 2 and “≤15 years” to 3), and 3) tobacco smoking after puberty or >15 years (“yes” to 1 and/or 2 and “>15 years” to 3).

Timing of body shapes (BS): Measured using a validated figural drawing scale of nine sex-specific body shapes at three time points: age eight years, puberty (voice break/menarche) and at age 30 years (Supplementary Figure 1) (22, 23). We applied a cut-off of figure five or greater to classify overweight BS in males, and a cut-off of figure four or greater to classify overweight BS in females. We further classified parents’ BS status according to timing of overweight as 1) normal weight BS up to puberty (normal BS at age eight and at puberty; reference category), 2) overweight BS up to puberty (overweight BS at age eight and at puberty), and 3) overweight BS starting after puberty and measured at age 30 (overweight BS at 30). This tool has been validated against measured height and weight for current (22) and for past BS (23), and the cut-offs defined by ability to identify overweight BMI (25-30 kg/m^2^) (22).

### 2.3 Epigenetic age acceleration in offspring

DNA methylation in offspring was measured in DNA extracted from peripheral blood, using a simple salting out procedure. Bisulfite-conversion was undertaken using EZ96-DNAmethylation kits (ZymoResearch, Irvine, CA, USA) at the Oxford Genomics Centre (Oxford, UK), and methylation was assessed using Illumina Infinium MethylationEPIC Beadchip arrays (Illumina, Inc., CA, USA) with samples randomly distributed on microarrays to control against batch effects. Methylation intensity files were processed, and quality was assessed using minfi (24) using R version 4.4.1. The probe normalization and filters were conducted using ref-RCP from the ENmix package (25). Epigenetic predictor scores were generated by the methscore function (26) using Enmix, and by applying methylation beta value as an input.

Four epigenetic clocks were used: Horvath, GrimAge, PhenoAge and DunedinPACE clocks (13, 27–29). They were selected on the following premises: the Horvath clock, developed using samples from individuals of all ages and multiple tissues, may be a more accurate method for studying younger participants included in this study, and capture broader aspects of organismal aging (30). GrimAge, PhenoAge and DunedinPACE are newer clocks trained on health indicators and clinical biomarkers, making them more closely related to lifestyle factors and health outcomes (27–29). Whereas most epigenetic clocks estimate age in years at time of measurement, DunedinPACE measures the yearly pace, or ticking rate of ageing, and therefore has a different metric. Although the clocks are moderately correlated with each other, they were trained on different datasets and age-related CpGs and may therefore show varying associations with health outcomes and lifestyle factors (31). Principal component (PC) trained clocks were used for Horvath, GrimAge and PhenoAge as they have shown improved reliability and precision (32).

The primary outcome, *epigenetic age acceleration*, was derived as the residual from a linear regression of epigenetic age on chronological age, with positive values indicating accelerated epigenetic ageing and negative values indicating decelerated ageing (13).

### 2.4 Covariates

Offspring smoking was recorded in RHINESSA with the same questions as used in parents in RHINE/ECRHS. Offspring BMI was calculated based on measured height and weight. Grandparental education was used as a proxy for the parent’s childhood socioeconomic status (SES). SES was categorized as *middle/high* if at least one grandparent had completed university or high school, otherwise it was categorized as *low*.

### 2.5 Statistical analysis

Linear regression models were used to investigate the associations of parental i) smoking and ii) overweight body shape status, with offspring epigenetic age acceleration using the PCHorvath, PCGrimAge, DunedinPACE and PCPhenoAge epigenetic clocks. Each clock was treated as a separate outcome. The data had a hierarchical nature: offspring (level one), nested within families (level two; one father/mother may have several offspring), also nested within study centers (level three). Both models accounted for family and study center, either by using mixed-effect regression models or by computing robust standard errors using the R package jtool (33) (HC1 covariance matrix estimator). Analyses of offspring-mother pairs were conducted in a comparable manner.

Confounders and adjusting variables were identified using Directed Acyclic Graphs (DAGs) (Supplementary Figure 2). Both models were adjusted for offspring sex, chronological age, and parent’s childhood socioeconomic status (SES). In addition, offspring’s smoking was included as an adjusting variable in the parental smoking model, while offspring BMI was included in the parental overweight model.

The sample sizes in the paternal and maternal smoking models were sufficiently large to allow stratification by offspring sex across both smoking initiation time points. In contrast, the parental overweight model had too few observations in the category of overweight at age 30 (n=26), limiting sex-stratified analyses to parental overweight status up to puberty.

Sensitivity analyses were conducted in offspring-father pairs for both smoking and overweight models. First, offspring’s BMI was included as an additional adjusting variable in the father’s smoking model, and offspring’s smoking in the father’s overweight model, since these factors have been shown to influence a person’s epigenetic age (16, 18, 34, 35). Second, we ran analyses excluding offspring who had ever smoked in the father’s smoking analysis, and offspring with BMI ≥25 in the father’s overweight analyses. Third, regression analyses were restricted to adult offspring (≥18 years) as only the Horvath clock was trained across all ages (13).

We presented the estimates of associations as β coefficients (β) with 95% Confidence Intervals (95% CI). The analyses were performed using R, Core Team (2025). _R: A Language and Environment for Statistical Computing. R Foundation for Statistical Computing_, Vienna, Austria (www.R-project.org) and Stata 19 SE (StataCorp, College Station, TX, USA).

### 2.6 Data Availability

Data are available on reasonable request. Request for access to data can be directed to CS, the RHINESSA study Principal Investigator.

## 3. Results

### 3.1 General characteristics

In the smoking model (N=892, 52% male), 43% had parents who had never smoked, while 73% had never smoked themselves (Table 1). In the analysis of parents’ overweight (N=719, 52% male), 12% had parents with overweight body shape in childhood and puberty, 4% had parents with overweight body shape at age 30, and 33% were overweight themselves (BMI ≥25 kg/m^2^) (Table 1). Characteristics specified for the paternal and maternal lines are provided in Supplementary Tables 1 and 2.

**Table 1.** Characteristics of the study population (offspring) according to timing of parent’s smoking initiation and parent’s overweight body shapes (OW-BS)

| Characteristics | Timing of parent's smoking initiation |  |  | Timing of parents' overweight body shapes |  |  |
| --- | --- | --- | --- | --- | --- | --- |
|  | Never<br>(n=385) | ≤15 years<br>(n=124) | >15 years<br>(n=383) | Never<br>(n=609)<sup>b</sup> | OW-BS<br>≤puberty (n=84) | OW-BS at 30<br>(n=26) |
| Sex (male), % | 52 | 52 | 53 | 53 | 44 | 62 |
| Age, mean (SD) |  |  |  |  |  |  |
| Female | 27.0 (8.5) | 27.3 (7.8) | 26.7 (7.2) | 25.8 (8.0) | 28.2 (6.7) | 26.2 (7.9) |
| Male | 27.6 (8.1) | 28.1 (7.8) | 29.5 (8.1) | 27.6 (7.9) | 28.8 (7.4) | 18.4 (6.0) |
| Smoking, % |  |  |  |  |  |  |
| Never smoking | 73 | 64 | 61 | 58 | 58 | 58 |
| Former smoking | 14 | 21 | 21 | 20 | 18 | 12 |
| Current smoking | 13 | 15 | 18 | 12 | 17 | 12 |
| BMI (kg/m<sup>2</sup>), % |  |  |  |  |  |  |
| Low (<18) | 6.2 | 4.0 | 4.2 | 7.1 | 4.8 | 7.7 |
| Normal (18-24.9) | 62 | 58 | 57 | 56 | 52 | 50 |
| High (≥25) | 29 | 35 | 37 | 27 | 32 | 39 |
| Father's early life<br>SES, %<sup>a</sup> |  |  |  |  |  |  |
| Low | 19 | 23 | 27 | 52 | 39 | 58 |
| Middle/high | 44 | 49 | 46 | 26 | 30 | 23 |
| Unknown | 37 | 27 | 27 | 22 | 31 | 19 |

Abbreviations: OW-BS= overweight body shapes; BMI= Body Mass Index; SES= Socioeconomic status Missing values in smoking analysis: smoking (n=1), BMI (n=20), parental childhood SES. Missing values in body shape analysis: smoking (n=70), BMI (n=71), parental childhood SES (n=167). <sup>a</sup> Grandparent educational level, <sup>b</sup> Never before conception

**Table 2.**
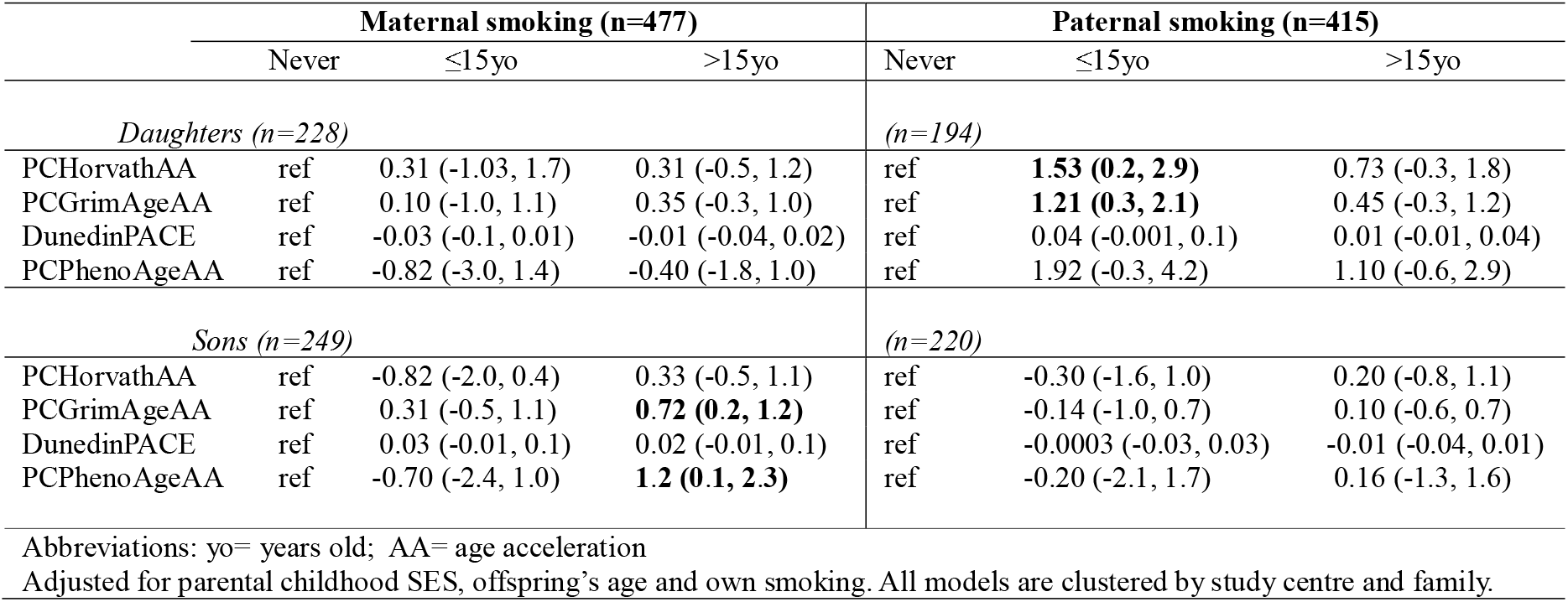
Parental smoking before or during puberty and after puberty, as associated with epigenetic age acceleration (AA) in daughters and sons (β [95% CI])

### 3.2 Accelerated epigenetic age in daughters of fathers starting smoking in puberty

Daughters whose fathers started smoking in puberty had consistently higher epigenetic age acceleration across all epigenetic clocks, statistically significant for PCHorvath (β 1.53; 95%CI 0.2, 2.9) and PCGrimAge (β 1.21; 95%CI 0.3, 2.1), and the age acceleration appeared to be higher than if the father started smoking after puberty (Table 2). No effect was indicated for daughters in relation to mother’s puberty smoking onset (PCHorvath [β 0.31; 95%CI −1.03, 1.7] and PCGrimAge [β 0.10; 95%CI −1.0, 1.1]), or in sons in relation to puberty smoking onset in either parent. Sons of mothers who started smoking after puberty had increased PCHorvath and PCPhenoAge acceleration (Table 2).

### 3.3 Accelerated epigenetic age in offspring of fathers who were overweight in puberty

If the father had been overweight in childhood and pubertal years, both daughters and sons showed consistently higher age acceleration, most pronounced in sons (PCHorvath [β 3.10; 95%CI 1.6, 4.6] and PCGrimAge [β 2.58; 95%CI 0.4, 4.8]) (Table 3). Comparable effects were not identified in relation to mother’s early onset overweight (sons: PCHorvath [β −1.03; 95%CI −2.2, 0.09] and PCGrimAge [β 0.12; 95%CI −0.6, 0.9]). In pooled analysis of parents’ overweight onset in adulthood in addition to childhood/puberty (Table 4), father’s overweight at 30 years showed no clear association with offspring’s epigenetic age acceleration, contrary to the consistent associations observed if father’s overweight started in childhood/puberty. Mother’s overweight at either age period showed no consistent associations with offspring epigenetic age acceleration.

**Table 3.** Parental overweight body shape (OW-BS) before or during puberty and after puberty, as associated with epigenetic age acceleration (AA) in daughters and sons (β [95% CI])

| Maternal overweight body shape (n=381) |  |  |  | Paternal overweight body shape(n=338) |  |  |
| --- | --- | --- | --- | --- | --- | --- |
| | Never<sup>a</sup> | OW-BS ≤puberty | OW-BS 30yo | Never<sup>a</sup> | OW-BS ≤puberty | OW-BS 30yo |
| *Daughters (n=187)* |  |  |  | *(n=159)* |  |  |
| PCHorvathAA | ref | 0.66 (-0.8, 2.1) | .. | ref | **1.35 (0.2, 2.6)** | .. |
| PCGrimAgeAA | ref | 0.57 (-0.5, 1.7) | .. | ref | -0.03 (-2.1, 2.0) | .. |
| DunedinPACE | ref | -0.02 (-0.1, 0.02) | .. | ref | 0.04 (-0.03, 0.1) | .. |
| PCPhenoAgeAA | ref | 2.02 (-0.2, 4.2) | .. | ref | **2.43 (0.5, 4.4)** | .. |
| *Sons (n=194)* |  |  |  | *(n=179)* |  |  |
| PCHorvathAA | ref | -1.03 (-2.2, 0.1) | .. | ref | **3.10 (1.6, 4.6)** | .. |
| PCGrimAgeAA | ref | 0.12 (-0.6, 0.9) | .. | ref | **2.58 (0.4, 4.8)** | .. |
| DunedinPACE | ref | -0.03 (-0.1, 0.01) | .. | ref | **0.11 (0.02, 0.2)** | .. |
| PCPhenoAgeAA | ref | -0.88 (-2.4, 0.7) | .. | ref | **3.30 (1.2, 5.4)** | .. |

Abbreviations: OW-BS= overweight body shape; AA= age acceleration; yo= years old; <sup>a</sup> = never before conception. Adjusted for parental childhood SES, offspring's age and BMI. All models are clustered by study centre and family.

**Table 4.** Parental overweight body shape (OW-BS) before and during puberty and after puberty - at age 30 years, as associated with epigenetic age acceleration (AA) in offspring. Sons and daughters are analyzed together in order to assess potential differences between time windows (β [95% CI])

| Maternal overweight body shape (n=381) |  |  |  | Paternal overweight body shape (n=338) |  |  |
| --- | --- | --- | --- | --- | --- | --- |
| | Normal | OW-BS ≤puberty | OW-BS 30yo | Normal | OW-BS ≤puberty | OW-BS 30yo |
| PCHorvathAA | ref | -0.08 (-1.0, 0.8) | **-1.61 (-3.1, -0.2)** | ref | **2.25 (1.2, 3.3)** | 0.41 (-1.2, 2.0) |
| PCGrimAgeAA | ref | 0.31 (-0.4, 1.0) | -0.51 (-1.8, 0.8) | ref | 1.36 (-0.2, 2.9) | 0.57 (-0.6, 1.7) |
| DunedinPACE | ref | -0.03 (-0.1, 0.002) | -0.04 (-0.1, 0.02) | ref | **0.07 (0.01, 0.1)** | -0.02 (-0.1, 0.03) |
| PCPhenoAgeAA | ref | 0.62 (-0.7, 2.0) | -1.72 (-4.6, 1.2) | ref | **3.11 (1.9, 4.4)** | 1.37 (-0.5, 3.3) |

Abbreviations: OW-BS= overweight body shape; AA= age acceleration; yo= years old. Adjusted for parental childhood SES, offspring's sex, age and BMI. All models are clustered by study centre and family.

### 3.4 Sensitivity analyses

When adjusting for offspring BMI and excluding ever-smoking offspring, a consistent faster age acceleration was found among daughters whose father’s started smoking in puberty (Supplementary Table 3). Similarly, when excluding offspring under age18, daughters whose fathers started smoking in puberty tended to have faster age acceleration (Supplementary Table 3). As for the analyses of fathers’ overweight up to puberty, effect estimates showed faster age acceleration in most epigenetic clocks when adjusting for offspring own smoking and when excluding offspring who were overweight themselves (BMI<25 kg/m^2^) (Supplementary Table 4). Similarly, analysis excluding offspring under age 18, showed slightly attenuated, but similar direction of effects as in the main model (Supplementary Table 4).

## 4. Discussion

Our findings provide evidence suggesting epigenetic ageing may result from exposures occurring in the parent generation and, notably, that father’s exposures before puberty may influence biological ageing in the next generation, independent of the offspring’s own lifestyle factors. Specifically, we observed accelerated epigenetic ageing in young and adult offspring of fathers who smoked (a chemical exposure) or were overweight (a metabolic exposure) in his puberty, whereas no associations were seen when these exposures occurred later, or if the mother was overweight or smoked in her puberty. The analyses of both fathers’ smoking and of father’s overweight showed consistent results when adjusting for the offspring’s own smoking and BMI, respectively, and in sensitivity analyses excluding offspring who had ever smoked or were overweight themselves. Epigenetic clocks, age-related variations in DNA methylation (the addition of methyl groups onto specific DNA sites), are biological age estimators that we used to calculate offsprings epigenetic age and epigenetic age acceleration (i.e. having an epigenetic age that exceed the chronological age). To our knowledge, this is the first analysis linking biological ageing in youngsters and adults to two different and important factors of their father’s lifestyle and environment when he was in puberty.

A critical developmental window in male puberty in humans has previously been suggested in analyses of father’s exposures in relation to specific health outcomes in offspring (2, 3, 5, 6, 9, 36, 37), and in a recent analysis of father’s adolescent smoking and offspring cord blood epigenetic age (20). Epidemiological reports from the ECRHS/RHINE/RHINESSA cohorts as well as the Tasmanian Longitudinal Health Study (TAHS) consistently show that paternal adolescent smoking and overweight are associated with offspring asthma and lung function (2-5, 36, 38, 39). In previous epigenome-wide association studies, we showed that both fathers’ early adolescent smoking exposures as well as being overweight around puberty, were associated with unique altered epigenetic patterns in the RHINESSA offspring, which were further linked to respiratory- and BMI-related outcomes (11, 12).

Notably, clear sex-specific patterns were found for fathers’ smoking in associations with offspring epigenetic ageing. The results were limited to daughters, but not sons, who had a consistently increased PCHorvath and PCGrimAge acceleration if the father had smoked in puberty. Fathers’ overweight body shape before puberty, on the other hand, was associated with increased PCHorvath and PCPhenoAge acceleration in both sons and daughters, yet stratified analyses indicated this pattern to be more pronounced and consistent in the sons, with accelerated aging across all the epigenetic clocks tested. Likewise, preconception paternal smoking has previously been linked to increased asthma risk (40) and BMI (41) in daughters, while paternal obesity has been shown to affect the phenotypes of his sons, particularly with respect to fetal development and risk of metabolic disorders and cardiovascular disease (42). Conversely, a study of paternal adolescent smoking and age acceleration in offspring cord blood, found the effects to be more pronounced in male newborns (20). One may suspect that differences in sex specificity could be related to differences in susceptibility to environmental stressors during early life development or in DNA methylation signals by fetal sex across perinatal tissues (20). However, there are several mechanisms that may lead to different sex-specific patterns in intergenerational studies (43).

Regarding the maternal line, a later exposure time period - maternal smoking initiation after puberty - was related to offspring epigenetic age acceleration only in sons and may reflect the consequences of *in utero* exposure. In line with these findings, a previous study from The Accessible Resource for Integrated Epigenomic Studies (ARIES), found that maternal BMI and smoking during pregnancy were correlated with higher epigenetic age acceleration in children (19). Although associations with maternal BMI tended to resolve during childhood, age acceleration in offspring of smoking mothers became larger during childhood and adolescence.

Several studies have demonstrated that a person’s epigenetic age is reflective of exposures during one’s own lifespan, and multiple measures of age acceleration have been found to be associated with obesity, both in adolescents and adults (18, 35, 44). A recent study show causal relationships between obesity and acceleration of epigenetic ageing (45). It has been suggested that obesity modifies DNA methylation patterns associated with biological ageing in different tissues and can increase the likelihood of aging-related disease risks (46).

Exposure to smoking has also been shown to accelerate epigenetic ageing both in the lungs and blood (16, 34, 47), with further impact on respiratory health and development of pulmonary diseases (16). The present study adds importantly to the understanding of risk factors for biological ageing by showing that also *fathers’* pubertal smoking and overweight appear to influence epigenetic age acceleration. Whether accelerated epigenetic ageing may be an underlying molecular mechanism and a potential link between paternal preconception exposures and offspring respiratory and metabolic health, is left to conjecture, and needs to be further investigated. As such, this study should be considered exploratory research, and a direction for future work.

A key strength of the study is the use of two linked intergenerational cohorts, enabling investigation of exposure-outcomes across generations based on parental data provided by the parents themselves and offspring data by the offspring themselves. Another main strength is the availability of data that allowed specifying the timing of two different pubertal exposures in both fathers and mothers and relating these preconception smoking and overweight periods to extensive data in their offspring, including multiple measures of epigenetic age acceleration. Moreover, the offspring span a wide age range and originated from different study centers, representing a heterogeneous group. For this reason, all analyses were clustered by study center and adjusted for offspring age. Since only the Horvath epigenetic clock was trained on both children and adults (13), we additionally ran sensitivity analyses restricted to adult offspring. In the analyses of fathers’ body shape status up to puberty, effect estimates for most clocks were only slightly attenuated after excluding offspring under 18 years. For the paternal smoking model, estimates among adult daughters whose fathers started smoking during puberty showed the same trend towards accelerated epigenetic ageing.

A potential limitation of the study is that different sets of offspring were included in the analyses comparing fathers and mothers. Although the offspring did not differ substantially in age, sex or other demographic and anthropometric data, we cannot rule out that the results may have been influenced by the difference in samples. The limited number of offspring with parental overweight at age 30 prohibited sex specific analyses for this aspect, otherwise, sample sizes were adequate. Information on paternal smoking and body shapes was based on self-reports, however, validation studies found that self-reported data provided a valid and reliable tool for assessing smoking behaviors in cohort studies (48). The body shape tool has also been validated against measured height and weight at different time points (23). Recall bias due to self-report can, however, not be ruled out, but it is likely that bias in parental reporting of exposures is independent of offsprings measures of epigenetic age acceleration, resulting in non-differential bias attenuating rather than exaggerating observed results.

Phenotypic traits such as smoking and obesity are reflective of a complex combination of shared genetics (49), and a range of lifestyle factors (50). Biomarkers of epigenetic age acceleration have also been shown to be under the influence of single nucleotide polymorphisms and other genetic variants (51). Although we adjusted for the fathers’ early-life environment and additionally clustered by family to account for the potential impact of shared familiar environments, unmeasured factors related to social class such as nutrition, lifestyle habits and housing conditions, may potentially have influenced our findings.

## 5. Conclusion

This novel human study suggests that biological ageing may be influenced not only by exposures during an individual’s own life span but also those experienced by their father. Father’s puberty appears to be a critical susceptibility window with regard to ageing, since both a chemical exposure (smoking) and a measure of metabolic status (overweight) were associated with accelerated epigenetic ageing in offspring - only if these factors occurred around paternal puberty. There was no indication of a susceptibility window in mother’s puberty, and the difference between the paternal and maternal lines agrees with the well-known differences in male and female germ cell development.

A susceptible developmental window in male puberty with long-term implications for offspring health and ageing, has wide scientific and societal implications. Future research should explore generational effects of boys’ lifestyle and environment more broadly, underlying biological pathways, how this translates into susceptibility to age-related diseases, and mechanisms that open for intervention, mitigation or adaption to such ancestral disadvantage.

## Data Availability

All data produced in the present study are available upon reasonable request to the authors

## Authors’ contributions (CRediT)

JPLC and TMØ contributed equally as joint first authors; CS and JWH contributed equally as joint last authors. Conceptualization: JPLC, TMØ, NTK, ML, RJB, SA, CJ, SCD, KAF, FJCG, MH, AJ, CL, AM, AO, FGR, AFV, VS, CS, JWH. Data curation: all authors. Formal analysis: JPLC, TMØ, NTK, CS. Funding acquisition: RJB, CJ, SCD, KAF, FJCG, MH, AM, AJ, CL, AO, FGR, VS, CS, JWH. Investigation: all authors. Methodology: all authors. Project administration: all authors. Writing - original draft: JPLC, TMØ, NTK, CS, JWH. Writing - review & editing: all authors.

## Acknowledgments

All co-authors declare no conflict of interest. Co-ordination of the RHINESSA study has received funding from the Research Council of Norway (Grants Nos. 274767, 214123, 228174, 230827 and 273838), ERC StG project BRuSH #804199, the European Union’s Horizon 2020 research and innovation programme under grant agreement No. 633212 (the ALEC Study), the Bergen Medical Research Foundation, and the Western Norwegian Regional Health Authorities (Grants Nos. 912011, 911892 and 911631). Study centres have received local funding from the following: Bergen: the above grants for study establishment and co-ordination, and, in addition, World University Network (REF and Sustainability grants), Norwegian Labour Inspection, and the Norwegian Asthma and Allergy Association. Albacete and Huelva: Sociedad Española de Patología Respiratoria (SEPAR) Fondo de Investigación Sanitaria (FIS PS09). Gøteborg, Umeå and Uppsala: the Swedish Heart and Lung Foundation, the Swedish Asthma and Allergy Association. Reykjavik: Iceland University. Melbourne: National Health and Medical Research Council (NHMRC) of Australia (research grants 299901 and 1021275). Tartu: The Estonian Research Council (Grant No. PUT562). Århus: The Danish Wood Foundation (Grant No. 444508795), the Danish Working Environment Authority (Grant No. 20150067134), Aarhus University (Ph.D. scholarship).

