## Supplementary Figure 1 for "Epigenetic age acceleration in offspring linked to paternal smoking initiation and overweight in puberty: Evidence from a two-generation study"

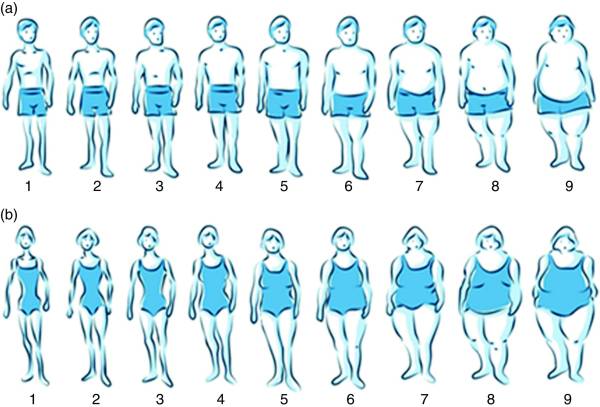

Footnote: Designed for ECRHS III / RHINE III by Alejandro Villén-Real

Legend Supplementary Figure 1. Figural drawing scales of the body shape tool

for a) men and b) women
