## Supplementary Figure 2 for "Epigenetic age acceleration in offspring linked to paternal smoking initiation and overweight in puberty: Evidence from a two-generation study"

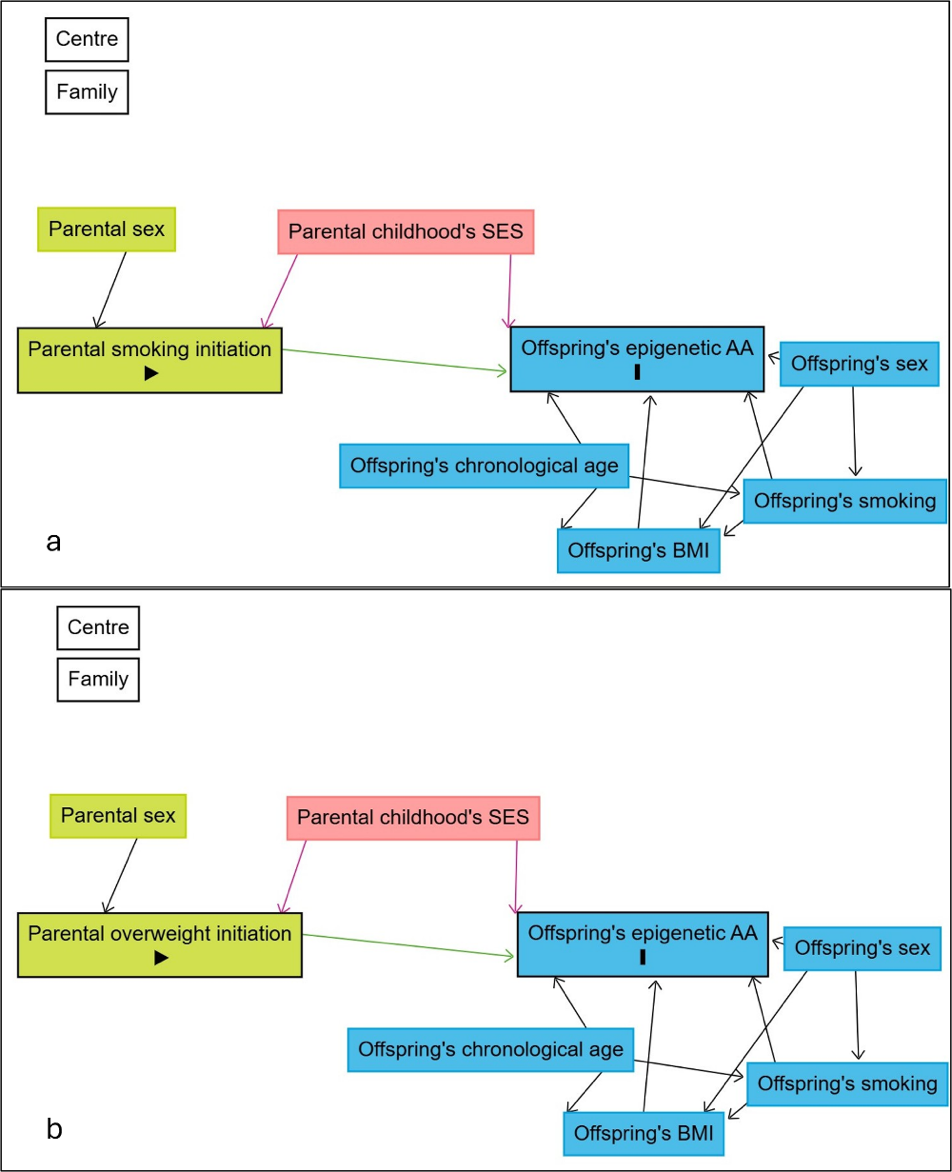

**Footnote:**
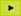
 exposure;
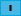
 outcome;
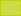
 ancestor of exposure;
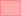
 ancestor of exposure and outcome;
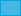
 ancestor of outcome;
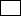
 other variable;
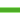
 causal path;
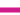
 biasing path.

Center and Family represent clustering variables used to account for non-independence in the data; they are not assumed to have a direct causal relation with the exposure or outcome.

Legend Supplementary Figure 2. Direct acyclic graphs (DAGs) of the

relationships between a) parental smoking initiation and

offspring’s epigenetic age acceleration (AA), and b) parental

overweight initiation and offspring’s epigenetic age AA.
