## Supplementary Table 1 for "Epigenetic age acceleration in offspring linked to paternal smoking initiation and overweight in puberty: Evidence from a two-generation study"

Supplementary Table 1. Characteristics of the study population (offspring) according to timing of father’s smoking initiation and overweight body shapes (OW-BS) (β [95% CI])

|  | **Timing of father’s smoking initiation** | | | **Timing of father’s overweight body shape** | | |
| --- | --- | --- | --- | --- | --- | --- |
| **Characteristics** | **Never (n=164)** | **≤15 years (n=67)** | **>15 years**  **(n=184)** | **Never (n=312) ^b^** | **OW-BS ≤puberty (n=20)** | **OW-BS at 30 (n=6)** |
| Sex (male), % | 52 | 52 | 54 | 54 | 40 | 50 |
| Age, mean (SD) | |  |  |  |  |  |
| Female | 25.1 (8.4) | 25.8 (7.5) | 26.8 (6.3) | 24.9 (7.4) | 27.7 (6.6) | 23.3 (6.5) |
| Male | 26.1 (8.8) | 27.2 (7.4) | 29.5 (7.9) | 27.0 (8.2) | 31.3 (8.0) | 15.7 (2.1) |
| Smoking, % |  |  |  |  |  |  |
| Never smoking | 79 | 70 | 62 | 59 | 35 | 33 |
| Former smoking | 10.4 | 15 | 21 | 13 | 20 | 17 |
| Current smoking | 10.4 | 15 | 17 | 11 | 35 | ·· |
| BMI (kg/m^2^), % |  |  |  |  |  |  |
| Low (<18) | 7.1 | 6.2 | 3.3 | 7.7 | 5.0 | ·· |
| Normal (18-24·9) | 66 | 55 | 57 | 55 | 45 | 83 |
| High (≥25) | 27 | 39 | 40 | 28 | 35 | 17 |
| Father’s early SES, %^a^ | |  |  |  |  |  |
| Low | 19 | 19 | 27 | 55 | 30 | 67 |
| Middle/high  Unknown | 46  35 | 46  34 | 45  29 | 24  21 | 15  55 | 33  ·· |
| Abbreviations: OW-BS= overweight body shapes; BMI= Body Mass Index; SES= Socioeconomic status  Missing values: in smoking analysis: Offspring’s smoking (n=1), BMI (n=13); in body shape analysis: offspring’s smoking (n=60), BMI (n=32). ^a^ Grandparent educational level. ^b^ Never before conception | | | | | | |
