## Supplementary Table 2 for "Epigenetic age acceleration in offspring linked to paternal smoking initiation and overweight in puberty: Evidence from a two-generation study"

Supplementary Table 2. Characteristics of the study population (offspring) according to timing of mother’s smoking initiation and overweight body shapes (OW-BS) (β [95% CI])

| **Timing of mother’s smoking initiation** | | | | **Timing of mother’s overweight body shape** | | |
| --- | --- | --- | --- | --- | --- | --- |
| **Characteristics** | **Never (n=221)** | **≤15 years (n=57)** | **>15 years (n=199)** | **Never (n=297) ^b^** | **OW-BS ≤puberty (n=64)** | **OW-BS at 30 (n=20)** |
| Sex (male), % | 52 | 53 | 52 | 51 | 45 | 65 |
| Age, mean (SD) | |  |  |  |  |  |
| Female | 28.4 (8.3) | 29.1 (8.0) | 26.6 (7.9) | 26.7 (8.5) | 28.4 (6.8) | 27.4 (8.6) |
| Male | 28.6 (7.4) | 29.1 (8.2) | 29.5 (8.3) | 28.4 (7.5) | 28.1 (7.3) | 19.1 (6.6) |
| Smoking, % |  |  |  |  |  |  |
| Never smoking | 69 | 58 | 59 | 50 | 63 | 50 |
| Former smoking | 16 | 28 | 21 | 19 | 17 | 5.0 |
| Current smoking | 15 | 14 | 20 | 12 | 11 | 5.0 |
| BMI (kg/m^2^), % |  |  |  |  |  |  |
| Low (<18) | 6.0 | 2.0 | 5.0 | 56 | 55 | 40 |
| Normal (18-24·9) | 62 | 63 | 58 | 27 | 31 | 45 |
| High (≥25) | 31 | 33 | 35 | 6.4 | 4.7 | 10 |
| Father’s early SES, %^a^ | |  |  |  |  |  |
| Low | 19 | 28 | 28 | 48 | 42 | 55 |
| Middle/high  Unknown | 43  38 | 53  19 | 47  25 | 29  23 | 34  23 | 20  25 |
| Abbreviations: OW-BS= overweight body shapes; BMI= Body Mass Index; SES= Socioeconomic status  Missing values: in smoking analysis: BMI (n=7); in body silhouette analysis: offspring’s smoking (n=73), BMI (n=39). ^a^ Grandparent educational level. ^b^ Never before conception | | | | | | |
