## Supplementary Table 3 for "Epigenetic age acceleration in offspring linked to paternal smoking initiation and overweight in puberty: Evidence from a two-generation study"

Supplementary Table 3. Sensitivity analyses of association between fathers smoking before

or during puberty and daughters epigenetic age acceleration (AA) in A) model with

additional adjustment for daughters BMI, B) a sub-sample of never smoking daughters,

C) a sub-sample of adult daughters (β [95% CI])

|  |  | Paternal smoking ≤15yo (n=142) | | |
| --- | --- | --- | --- | --- |
|  |  | A. Adjustment daughters BMI | B. Never smoking daughters | C. Adult daughters (≥18yo) |
| PCHorvathAA | ref | 1.10 (-0.4, 2.6) | 1.33 (-0.5, 3.2) | 0.70 (-0.9, 2.2) |
| PCGrimAA | ref | 1.00 (0.1, 1.9) | 1.13 (0.1, 2.2) | 0.78 (-0.1, 1.7) |
| DunedinPACE | ref | 0.04 (0.0004, 0.1) | 0.02 (-0.02, 0.1) | 0.03 (-0.01, 0.1) |
| PCPhenoAA | ref | 1.40 (-0.7, 3.5) | 1.61 (-1.0, 4.3) | 0.74 (-1.4, 2.9) |
| Abbreviations: AA= age acceleration; yo= years old. Model A. Adjusted for father’s childhood SES, offspring age, own smoking and offspring BMI, Model B. Adjusted for father’s childhood SES and offspring age, Model C. Adjusted for father’s childhood SES, offspring age and own smoking· All models clustered by study center and family origin | | | | |
