## Supplementary Table 4 for "Epigenetic age acceleration in offspring linked to paternal smoking initiation and overweight in puberty: Evidence from a two-generation study"

Supplementary Table 4. Sensitivity analyses of association between father’s overweight body shape (OW- BS) before and during puberty and offspring epigenetic age acceleration (AA) in A) model with additional adjustment for offspring smoking, B) a sub-sample of offspring with BMI<25 kg/m^2^, C) a sub-sample of adult offspring (β [95% CI])

|  | Paternal overweight body shape around puberty (n=361) | | | |
| --- | --- | --- | --- | --- |
|  |  | A. Adjustment offspring smoke | B. Offspring BMI<25 kg/m^2^ | C. Adult offspring (≥18yo) |
| PCHorvathAA | ref | 2.14 (1.2, 3.1) | 1.59 (-0.7, 3.9) | 2.21 (1.0, 3.4) |
| PCGrimAA | ref | 1.20 (-0.5, 2.9) | 1.74 (0.9, 2.6) | 1.30 (-0.4, 3.0) |
| DunedinPACE | ref | 0.07 (0.01, 0.1) | 0.10 (0.07, 0.1) | 0.07 (0.02, 1.1) |
| PCPhenoAA | ref | 2.74 (1.7, 3.8) | 1.09 (-0.4, 2.5) | 2.84 (1.7, 4.0) |
| Abbreviations: OW-BS= overweight body shape; AA= age acceleration; yo= years old; Model A. Adjusted for father’s childhood SES, offspring sex, age, BMI and own smoking, Model B. Adjusted for father’s childhood SES, offspring sex and age. Model C. Adjusted for father’s childhood SES, offspring sex, age and BMI. All models clustered by study center and family origin | | | | |
